# Predicting Health Disparities in Regions at Risk of Severe Illness to inform Healthcare Resource Allocations during Pandemics

**DOI:** 10.1101/2020.07.06.20147181

**Authors:** Tara Fusillo

**Affiliations:** John F. Kennedy, High School 3000 Bellmore Avenue, Bellmore, NY 11710

**Keywords:** Coronavirus, SARS-CoV-2, Pandemic, Socioeconomic status, Predictive model, Healthcare resource allocation

## Abstract

Pandemics including COVID-19 have disproportionately affected socioeconomically vulnerable populations. To create a repeatable modelling process to identify regional population centers with pandemic vulnerability, readily available COVID-19 and socioeconomic variable datasets were compiled, and linear regression models were built during the early days of the COVID-19 pandemic. The models were validated later in the pandemic timeline using actual COVID-19 mortality rates in states with high population densities, with New York, New Jersey, Connecticut, Massachusetts, Louisiana, Michigan and Pennsylvania showing the strongest predictive results. Our models have been shared with the Department of Health Commissioners of each of these states as input into a much needed “pandemic playbook” for local healthcare agencies in allocating medical testing and treatment resources.

## I. Introduction

Socioeconomic vulnerability can directly influence the severity of pandemics and their impact on mortalities in ways including access to healthcare, residence in crowded housing units and comorbidities. Prior studies of swine flu (H1N1) have pointed to these factors as contributors to the spread and severity of that pandemic^1^. Other studies have identified national level correlations that are helpful, but not actionable at a local level where actual healthcare resource allocation decisions are made^2^.

Early and accurate decisioning for healthcare resource allocations are particularly critical in geographic locations with high population density. This research created a repeatable modelling process that uses readily available data sources to identify the top counties in densely populated states with high likelihoods of disproportionate COVID-19 mortality rates. These models have been shared with the Department of Health Commissioners of New York, New Jersey, Connecticut, Massachusetts, Louisiana, Michigan and Pennsylvania where the models performed best. All Commissioners responded positively which validates our hypothesis that neither other researchers nor the Departments of Health themselves have developed a similar modelling process as a means to allocate scarce healthcare resources such as testing, treatment, education and communication.

## II. Methods

Exploratory data research at a national level was performed using county level data (Federal Information Processing Standards for county identification; also known as FIPS). COVID-19 mortality datasets (deaths per 100,000 of population) were created using the John’s Hopkins Dataset (https://coronavirus.jhu.edu/map.html)^3^ and data from the US Census Bureau^4^. Socioeconomic vulnerability datasets at a county level were created using sub-components of the CDC Social Vulnerability Index (https://svi.cdc.gov/data-and-tools-download.html) ^5^.

**Fig. 1.**
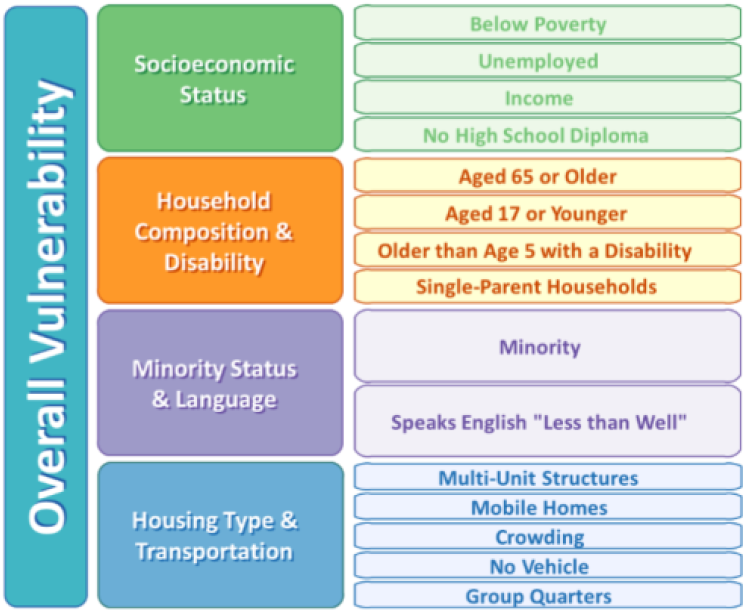
Full list of sub-component variables from the CDC (https://svi.cdc.gov/data-and-tools-download.html)

Scatterplots and trendlines were used to identify those variables most correlated with COVID-19 mortalities. Few, if any, social vulnerability variables correlated across all of the 3,142 FIPS counties, but minority status correlated strongly in certain regions, particularly those with high mortality rates.

**Fig. 2.**
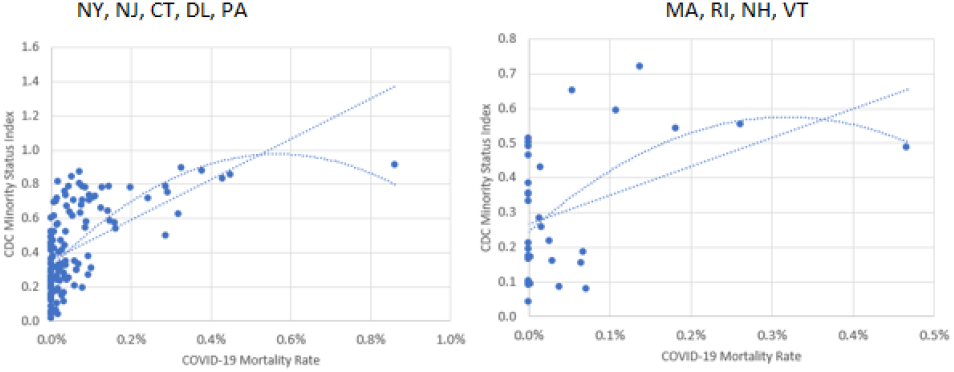
Examples of regional scatterplots and trendlines.

These findings focused the next steps on state level rather than national level correlations. County-level dependent variable datasets were created using COVID-19 mortality and population data from coronadatascraper.com (data service that scrapes county level COVID-19 data on a daily basis)^6^ as well as usafacts.org^7^ for cumulative mortalities as of April 8,2020 and May 8, 2020.

County-level independent variable datasets were created using socioeconomic data from countyhealthrankings.org, a collaboration dataset created by the Robert Wood Johnson Foundation and the University of Wisconsin Population Health Institute^8^. Given the year-to-year stability of most of these socioeconomic variables, the latest data available in the countyhealthrankings.org was used and no attempt was made to augment the dataset to try to match the time-series to either April 8, 2020 or May 8, 2020.

Cumulative COVID-19 specific mortality data (deaths per 100,000 of population) by county for high mortality rate states (NY, NJ, CT, MA, LA, MI, PA) as of May 8th was used as the dependent variable.

The May 8, 2020 date was used to ensure that the dependent variable would be tuned to a timeframe at or around the peak in daily mortalities when healthcare resources (testing, treatment, tracing) are typically most needed. The mortality curves below provide support for May 8th as the overall date for mortality predictions as shown in the Institute of Health Metrics and Evaluation dataset (https://covid19.healthdata.org/)^10^.

A stepwise linear regression technique was used to build each state level model. All relevant independent variables (Figure 3) were initially used in the model (i.e., include severe housing problems, but exclude violent deaths). Next, the variable with the lowest T-statistic was removed and the linear regression was re-run. This process was repeated until all T-statistics for the remaining independent variables were near a value of 2 or greater.

**Fig. 3.**
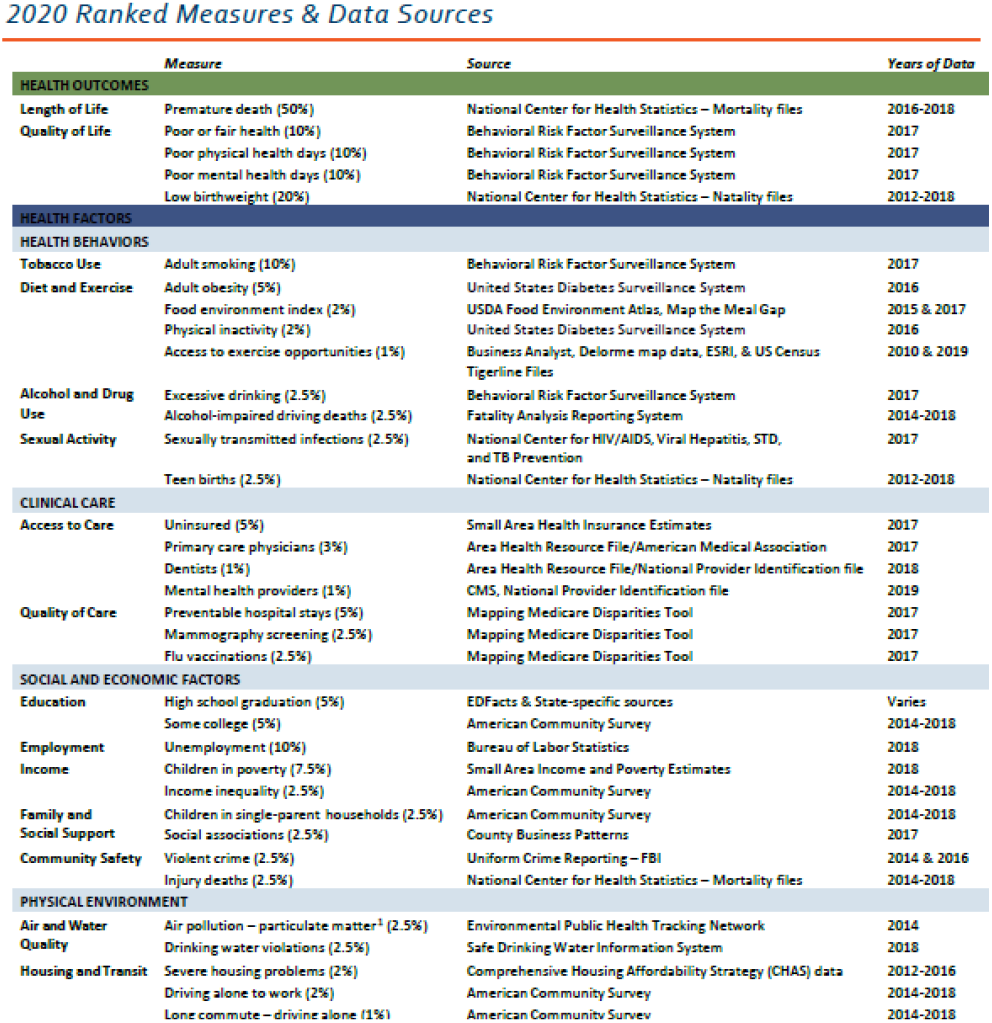
Full variable list from countyhealthrankings.org.

**Fig. 4.**
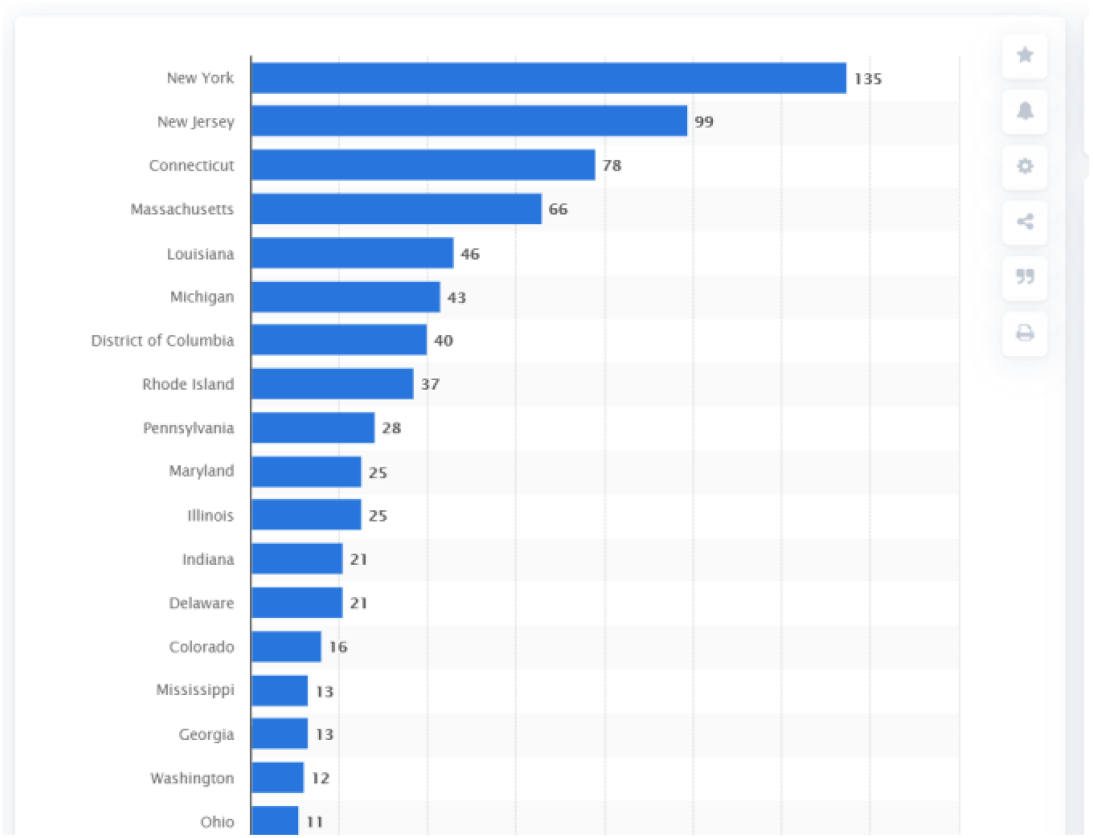
**COVID-19 mortality rates by state as of May 8, 2020 from statista.com^9^**

**Fig. 5.**
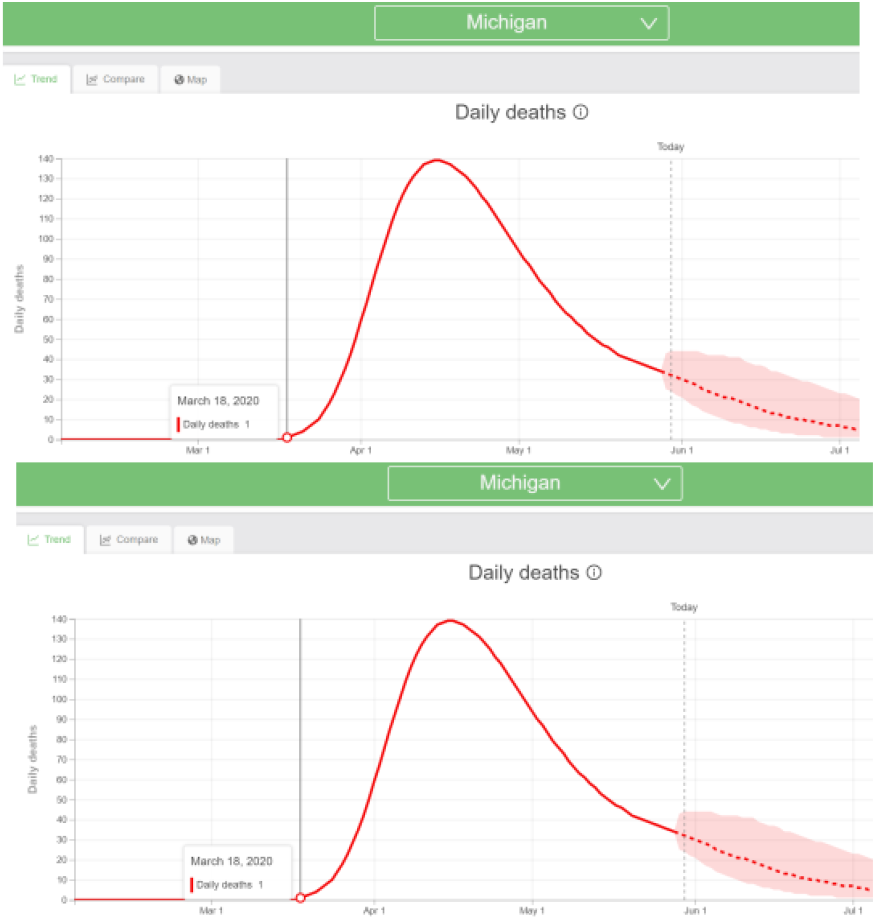
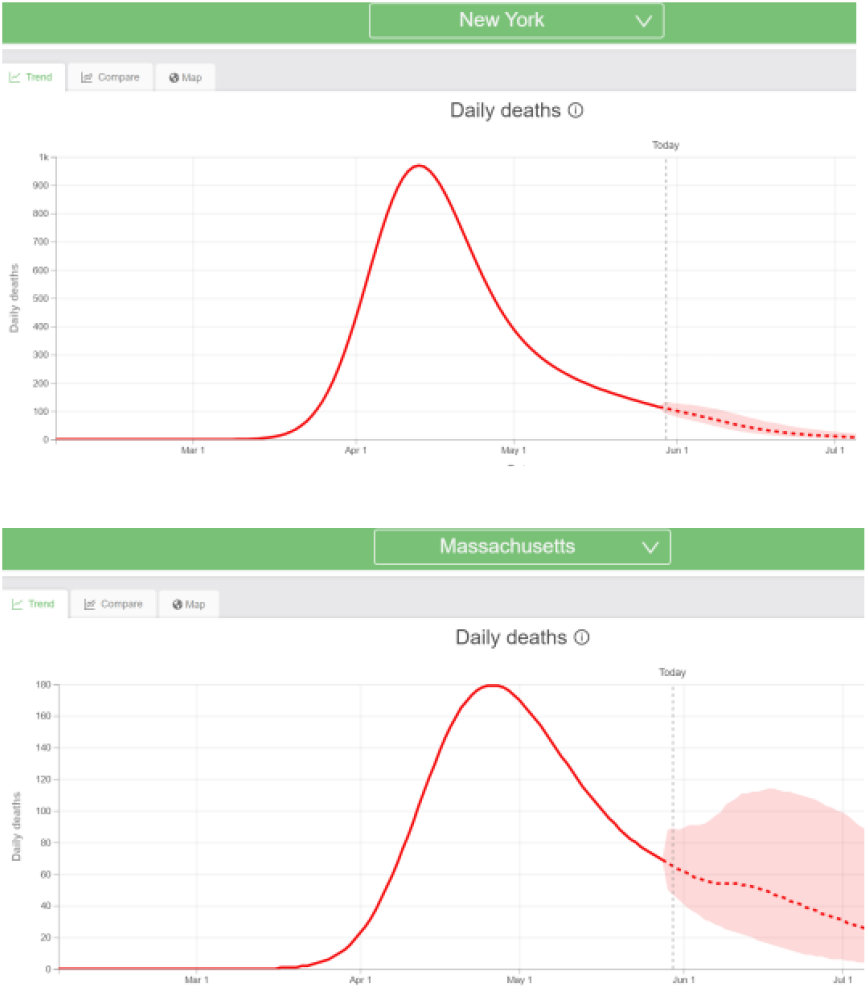
COVID-19 mortality curves examples from Institute of Health Metrics and Evaluation website (https://covid19.healthdata.org/)

## III. Results

Predictive models were completed for New York, New Jersey, Connecticut, Massachusetts, Louisiana, Michigan and Pennsylvania with statistically significant results.

**Fig. 6.**
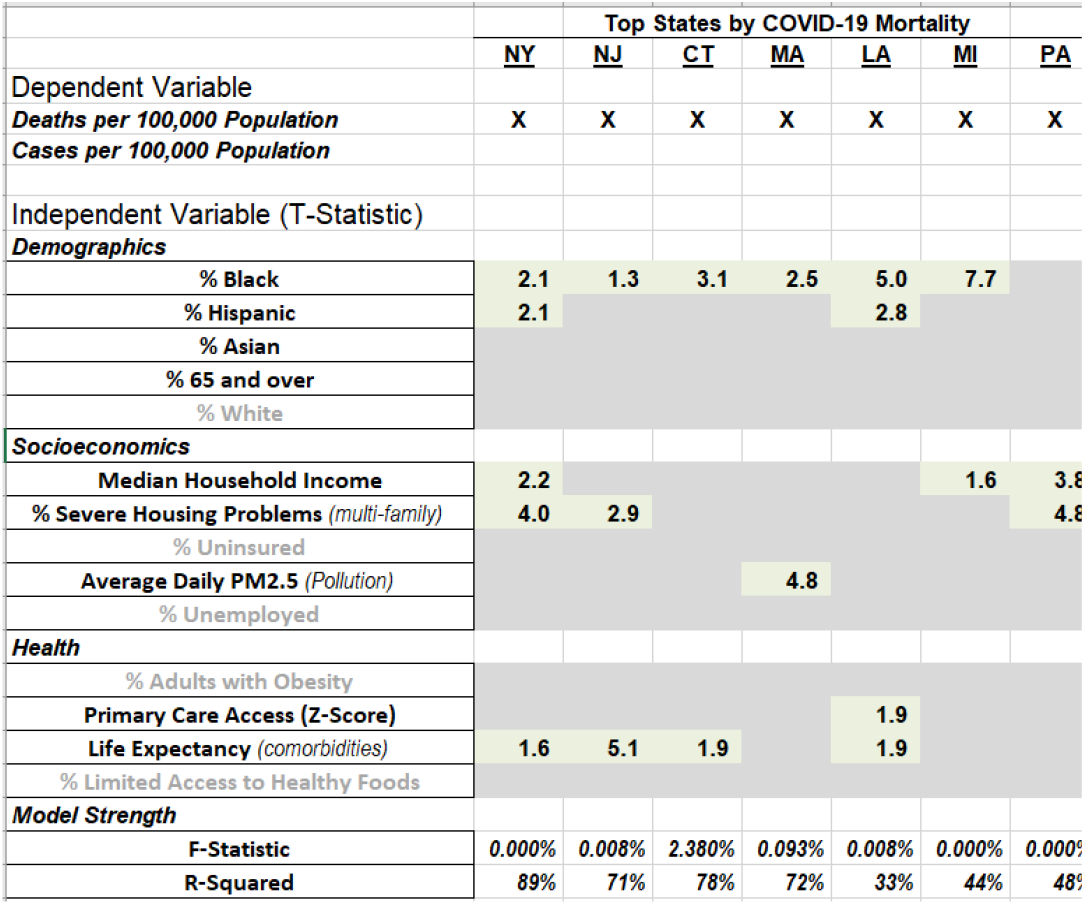
Final model variables and statistical significance.

Model validation comparing predicted to actual county rankings by cumulative mortality rate (deaths per 100,000 of population) through May 8^th^ and data visualizations using state level maps were completed. These validations were used to share model methods and results with the Commissioners of each state’s Department of Health.

**Fig. 7.**
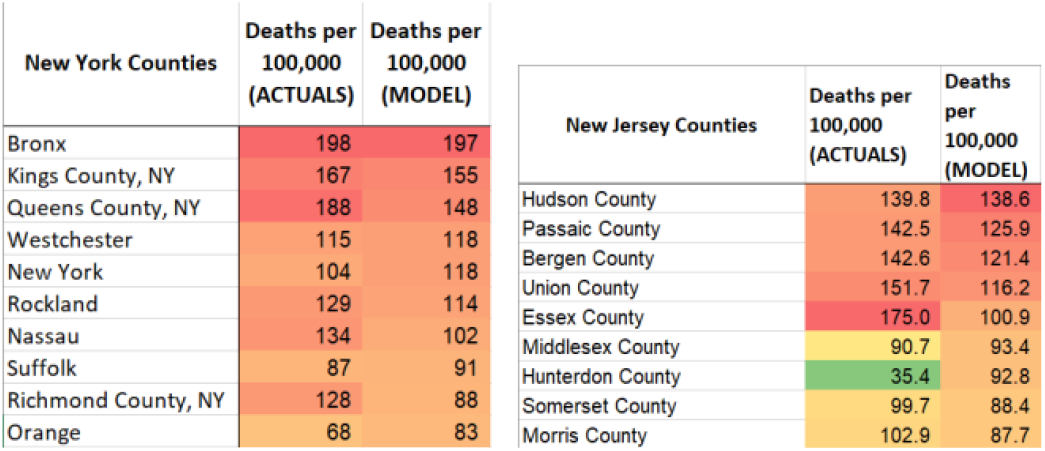
Top counties by mortality rate for New York and New Jersey (actuals vs. model prediction). Red indicates high mortality rates, yellow indicates medium and green indicates low relative to all other counties within a particular state

**Fig. 8.**
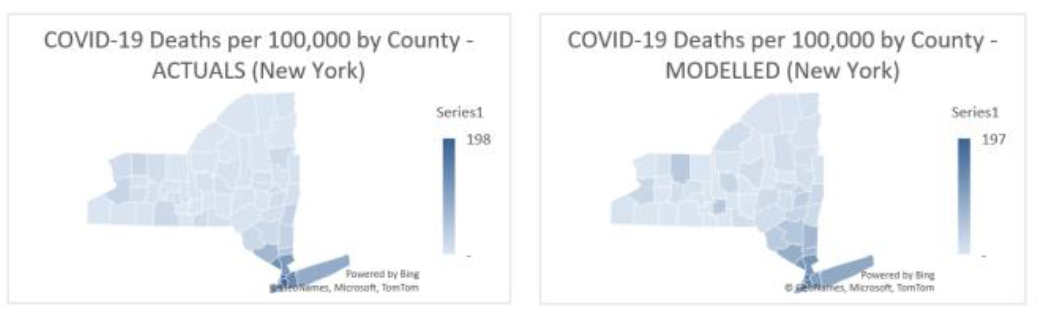
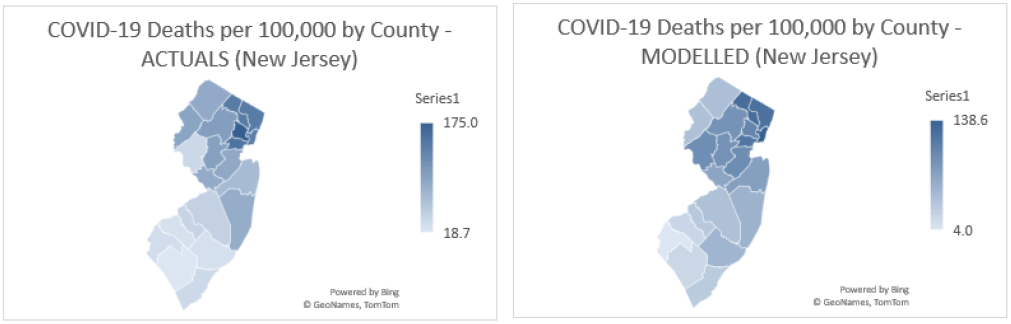
COVID-19 mortality rate heat maps for New York and New Jersey (actuals vs. model prediction)^11^.

Two further validations were completed as well. The first validation was to check model performance using COVID-19 mortality data on April 8, 2020 instead of May 8, 2020. This validation tested whether models using data available early in the pandemic would have been sufficient to build the models. The same variables were used, but coefficients were recalibrated with the April 8 dataset. April 8 and May 8 model outputs were compared to test for stability in the top counties predicted for high COVID-19 mortality rates. For New York, New Jersey and Connecticut, the models were stable. For Massachusetts, the 4/8 model performance was not strong, but this was easily corrected by using case data in place of mortality data. This is an important finding as it validates the predictive power contained in early case data which is more readily available at the start of a pandemic.

**Fig. 9.**
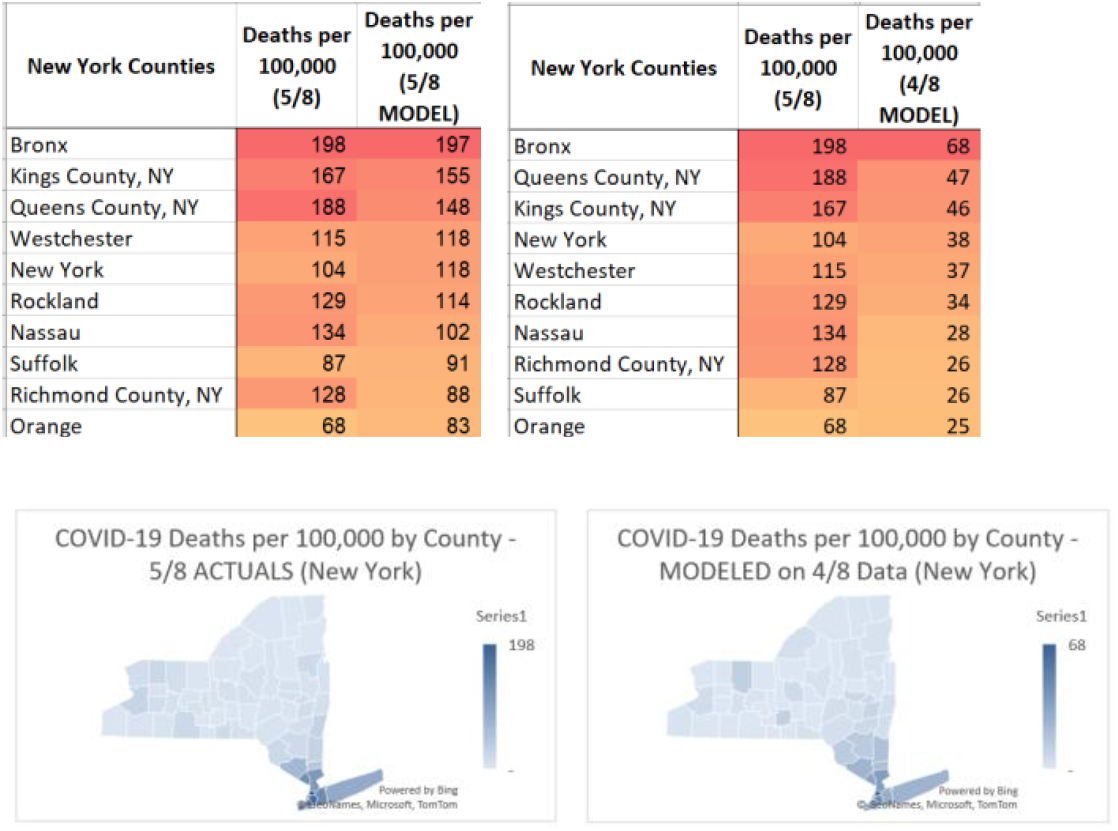
New York model validation results example using April 8 mortality data.

**Fig. 10.**
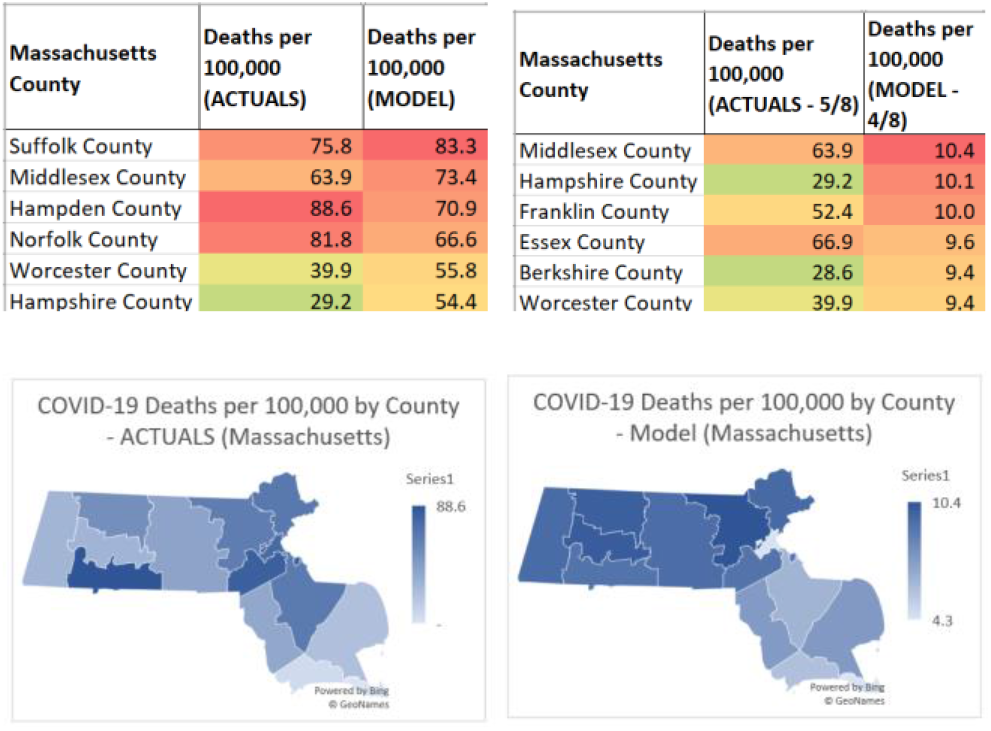
Massachusetts model validation results example using April 8 mortality data.

**Fig. 11.**
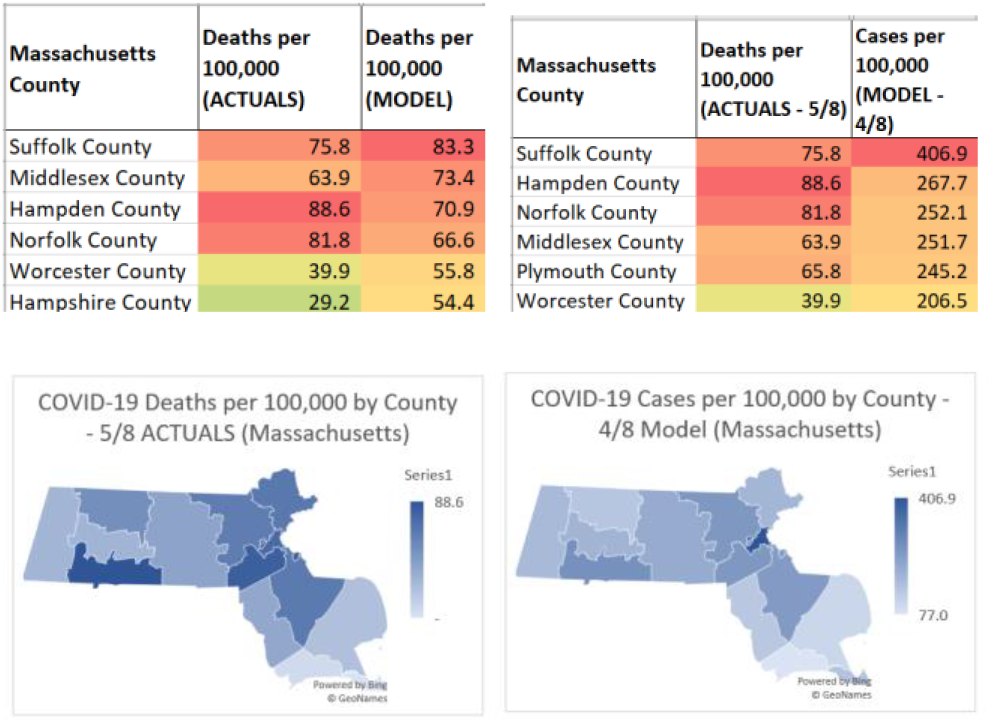
Massachusetts model validation results example using April 8 case data.

The second validation was to check the independent variables for multi-collinearity, with a specific focus on the correlations between Black and variables such as severe housing and uninsured. Strong multi-collinearity was seen in Connecticut, Massachusetts, Louisiana and Michigan partially explaining why these variables did not remain in the model after the stepwise regression process.

**Fig. 12.**
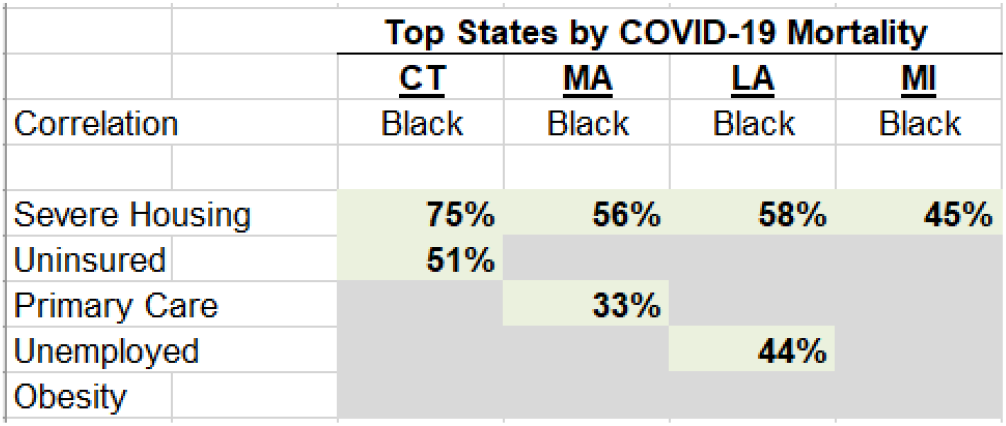
Multi-collinearity results for states without severe housing and other socioeconomically vulnerable variables.

With models and validations completed, the Departments of Health for New York, New Jersey, Connecticut, Massachusetts, Louisiana, Michigan and Pennsylvania were contacted. Additionally, agents of the New York Department of Health (Northwell Health and CORE) and a third-party statistical modelling firm for Connecticut (COVIDACTNOW) were contacted. Response from these contacts were positive and, in some cases, occurred within an hour of outreach (Northwell Health). This response indicates the strong appetite and need for this type of healthcare resource allocation tool for pandemics and other health crises. In fact, the Pennsylvania Department of Health indicated this tool’s importance in any second waves of COVID-19.

**Fig. 14.**
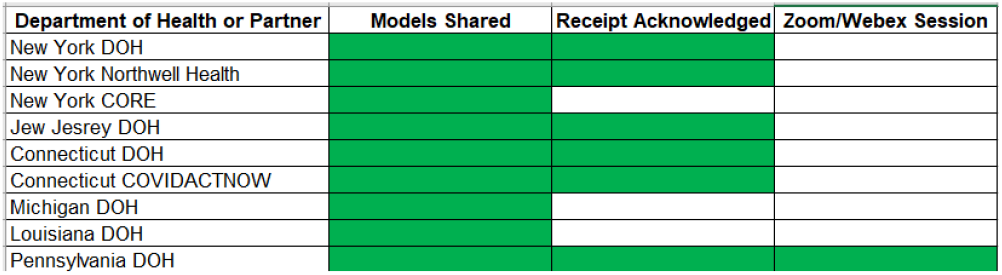
State Department of Health contact summary.

## IV. Discussion

The research described in this manuscript has shown the extent to which Black Americans, people living in crowded housing units, and households with less access to healthcare are at higher risk of severe illness during a pandemic. The results of other studies provide several possible explanations for these results. Lower income households are more likely to live in crowded housing units and multi-family homes^12^. Lower income households are also structurally disadvantaged in their access to medical insurance and healthcare^13^. Some studies have also pointed to a concept called “weathering” within the Black American community. Arline Geronimus, a professor of public health at the University of Michigan, showed through her research that among black communities, coping with financial strain, discrimination and barriers to good education elevates the stress response contributing to obesity, diabetes, hypertension and heart disease^14^.

The CDC conducted research on the co-occurrence of COVID-19 and certain ethnicities using a sample of 580 patients with lab-confirmed data^15^. Their results showed that more hospitalizations in Black patients than White patients. The researchers cited underlying medical conditions, work circumstance and living conditions to be major factors in COVID-19 mortalities in their sample. For example, members of racial and ethnic minorities are more likely to live in densely populated areas, making it more difficult to practice social distancing and being more susceptible to contracting and spreading COVID-19^15^. These members also lived farther away from grocery stores and medical facilities, thus being less able to receive necessary resources and medical attention^15^. Other examples cited included Hispanic and Black American workers employed in higher risk industries and often lacking paid sick leave. The researchers hypothesized that these types of workers were more likely to continue working despite being sick, thus exposing other workers to the disease^15^. The CDC recommended at the conclusion of this research that public health officials communicate to different population groups about COVID-19 and provide more healthcare services to ethnic minority groups^15^.

While studies such as these are insightful, we are not aware of any research that translates these impacts into predictive models that can be used locally to direct healthcare resources to the communities most likely to need them, thereby reducing mortalities caused by an ongoing set of institutional inequities.

That said, some organizations have attempted to create healthcare resource allocation methods using descriptive statistics. The Centers for Disease Control and Prevention (CDC) created the social vulnerability index (SVI), allowing the healthcare community to see which factors contribute to socioeconomic vulnerability. The CDC SVI factors are grouped into 4 groups: Socioeconomic status, Household Composition & Disability, Minority Status & Language, and Housing Type & Transportation. Although these factors are crucial inputs in identifying specific vulnerable communities, these four categories of factors alone are not sufficient to create the types of predictive models that state and local healthcare agencies can use. One example of this insufficiency is a recent study at Emory University where researchers identified correlations between COVID-19 mortalities and the SVI at a national level in the US^2^. While this study is helpful, it stopped short of recommending methods or processes to effectively distribute healthcare resources to specific counties in the US, particularly in the early days of a pandemic. Another study from the Surgo Foundation states that COVID-19 created new challenges for many communities with a dependence on health and structural factors, which are not completely captured by the CDC SVI^16^. In addition to the four socioeconomic factors provided by the CDC, the Surgo Foundation added two more factors: Epidemiologic Factors and Healthcare System Factors. Underlying health conditions in addition to healthcare system factors have been proven to greatly enhance a community’s vulnerability during a pandemic. The Surgo Foundation combined these two factors with the CDC SVI to create the COVID-19 Community Vulnerability Index (CCVI). The Surgo Foundation has created heatmaps to show retrospectively which counties were most vulnerable as measured by their CCVI. Similar to the Emory University study however, this methodology did not create a predictive model to identify where the mortalities would be highest at peak periods in a pandemic. We also compared the Surgo Foundation heatmap to our own predictive model rankings and confirmed that our projections of the top counties by per capita mortalities were far closer to reality.

**Fig. 14.**
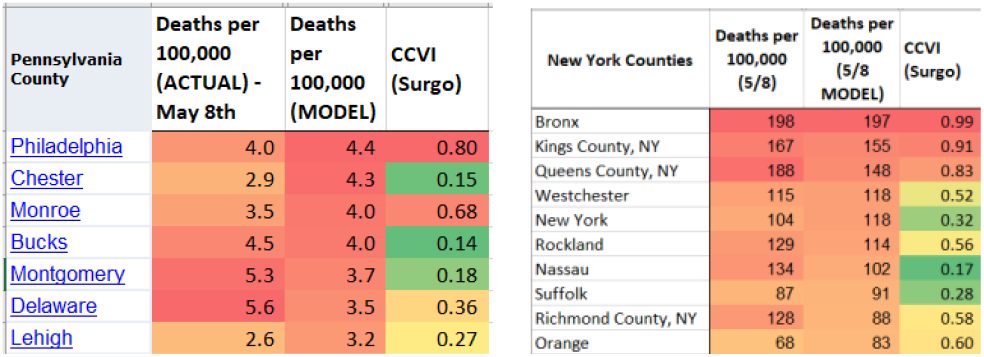
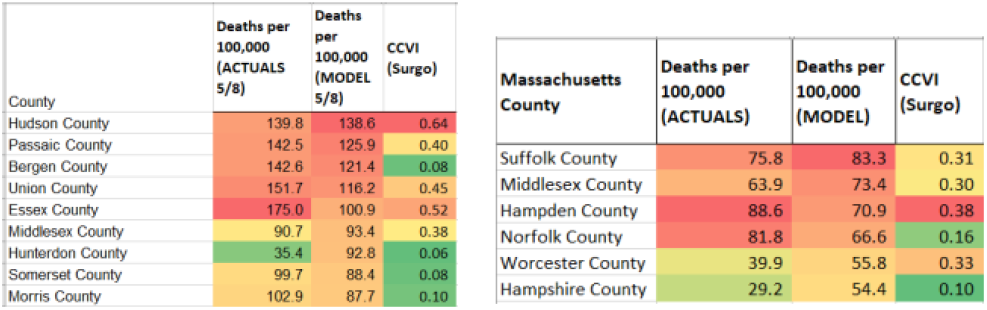
Comparison to Surgo Foundation Heatmap CCVI rankings.

During the early phases of the research described in this manuscript, we used the CDC’s SVI, similar to the Emory University study. We explored ***all*** of the sub-category factors in the SVI, but none showed strong correlations at a FIPS county level across the US. We then grouped states together by region and found strong correlations in the most densely populated regions, particularly with the minority status sub-factors. Similar to the Surgo Foundation study, we posited that the CDC’s SVI subfactors alone would be insufficient to build predictive models, so a far more complete independent variable dataset of socioeconomic and healthcare data was sourced from countyhealthrankings.org as discussed earlier. The key differentiation of our work are the predictive models ***for each state*** given the local differences in how each of the factors act as predictors of peak COVID-19 per capita mortalities. Our experience in meeting with the Pennsylvania Department of Health in early June 2020 confirms the uniqueness and usefulness of the approach given that they will be using the predictive modelling process for healthcare resource allocations in the event of a potential second wave of COVID-19 in the Fall of 2020. Other state level departments of health were similarly intrigued by our predictive approach including those in New York, New Jersey and Connecticut.

### Limitations

The models employed in the analyses are reliant on the accuracy of the datasets compiled. COVID-19 mortality data in particular has been notoriously difficult for states to report accurately at a county level throughout the pandemic for reasons including mortality cause classification errors at local coroner’s offices^17^.

## V. Conclusions

Our modelling process can be used to identify early in future pandemics or health crises, the communities most in need of healthcare resources. The COVID-19 models and the overall methodology have been received with enthusiasm by the New York, New Jersey, Connecticut, Massachusetts, Louisiana, Michigan and Pennsylvania Departments of Health.

## Data Availability

All data used in this manuscript are from publicly available sources.
[3]     COVID-19 Map. Johns Hopkins Coronavirus Resource Center, coronavirus.jhu.edu/map.html.
[4]     US Census Bureau, https://www.census.gov/data/tables/time-series/demo/popest/2010s-counties-total.html
[5]     CDC's Social Vulnerability Index (SVI). Centers for Disease Control and Prevention, Centers for Disease Control and Prevention, svi.cdc.gov/data-and-tools-download.html.
[6]     Davis, Larry. Corona Data Scraper, coronadatascraper.com/.
[7]     Coronavirus Live Map: US Coronavirus Cases by County. USAFacts, USAFacts, 23 June 2020, usafacts.org/visualizations/coronavirus-covid-19-spread-map/.
[8]     County Health Rankings, https://www.countyhealthrankings.org/
[9]     Elflein, John. Trailing Seven Day Average Number of COVID 19 Deaths Select Countries Worldwide 2020. Statista, 24 June 2020, www.statista.com/statistics/1111867/trailing-seven-day-average-number-of-covid-19-deaths-select-countries-worldwide/.
[10]    IHME: COVID-19 Projections. Institute for Health Metrics and Evaluation, covid19.healthdata.org/united-states-of-america.
[11]    Bing & TomTom data from the Excel mapping tools
● Tableau Dashboard on Vulnerability:   https://c19hcc.org/resource/vulnerable-population
● World Health Organization (WHO) dashboard: https://who.sprinklr.com/
● Full list of datasets: https://cgdv.github.io/challenges/COVID-19/datasource/
● Uninsured data: https://www.census.gov/data-tools/demo/sahie/#/data_2013
● COVID-19 deaths by ethnicity: https://www.apmresearchlab.org/covid/deaths-by-race
● FIPS to Zip code mapping: https://data.world/niccolley/us-zipcode-to-county-state/workspace/file?filename=ZIP-COUNTY-FIPS_2018-03.csv
● Nursing Home (Medicare) by Zip code: https://data.medicare.gov/data/nursing-home-compare
● Johns Hopkins tracking of states that currently provide COVID data by ethnicity (https://coronavirus.jhu.edu/data/racial-data-transparency)

## Acknowledgment

The author wishes to acknowledge Dr. Fathima Wakeel, Associate Professor at Lehigh University College of Health, for her mentorship throughout this project and for her review of this manuscript. Dr. Wakeel’s guidance was instrumental, particularly in directing the research toward a state level and in driving to actionable recommendations for state departments of health. The author also wishes to acknowledge Iwao Fusillo, Global Head of Data & Analytics for the National Football League (NFL) and Ms. Fusillo’s father, for helping her learn various techniques in dataset creation, statistical analysis and data visualization.

## Disclosures

The author has no conflicts of interest to disclose.

## Other datasets referenced in this research

- Tableau Dashboard onVulnerability: https://c19hcc.org/resource/vulnerable-population
- World Health Organization (WHO) dashboard: https://who.sprinklr.com/
- Full list of datasets: https://cgdv.github.io/challenges/COVID-19/datasource/
- Uninsured data: https://www.census.gov/data-tools/demo/sahie/#/data_2013
- COVID-19 deaths by ethnicity: https://www.apmresearchlab.org/covid/deaths-by-race
- FIPS to Zip code mapping: https://data.world/niccolley/us-zipcode-to-county-state/workspace/file?filename=ZIP-COUNTY-FIPS_2018-03.csv
- Nursing Home (Medicare) by Zip code: https://data.medicare.gov/data/nursing-home-compare
- Johns Hopkins tracking of states that currently provide COVID data by ethnicity (https://coronavirus.jhu.edu/data/racial-data-transparency)

## Other research papers evaluated during the course of this research

- Research paper based on China data (complicated paper, but some good ideas): https://doi.org/10.1101/2020.03.13.20035238doi
- Research paper on socio-economic status and cardio-vascular disease (not directly applicable to COVID-19, but interesting): https://www.ncbi.nlm.nih.gov/pmc/articles/PMC1694190/
- FastCompany – excellent article with many ideas for hypotheses to test: https://www.fastcompany.com/90479231/9-maps-that-show-which-areas-could-be-more-vulnerable-to-the-covid-19-pandemic
- Fivethirtyeight article: https://fivethirtyeight.com/features/the-young-americans-most-vulnerable-to-covid-19-are-people-of-color-and-the-working-class/
- The Conversation article: https://theconversation.com/covid-19-is-hitting-black-and-poor-communities-the-hardest-underscoring-fault-lines-in-access-and-care-for-those-on-margins-135615
- HealthAffairs article: https://www.healthaffairs.org/do/10.1377/hblog20200319.757883/full/
- Vox article: https://www.vox.com/identities/2020/4/7/21211849/coronavirus-black-americans
- FastCompany article on interesting data tool for Urban Footprint: https://www.fastcompany.com/90481120/this-tool-is-helping-cities-find-the-neighborhoods-most-vulnerable-to-coronavirus
- Persistent geographic variations in availability and quality of nursing home care in the United States: 1996 to 2016 | BMC Geriatrics: https://bmcgeriatr.biomedcentral.com/articles/10.1186/s12877-019-1117-z
- Do Lockdowns Save Many Lives? In Most Places, the Data Say No: https://www.wsj.com/articles/do-lockdowns-save-many-lives-is-most-places-the-data-say-no-11587930911?emailToken=f098d16b875951b5c8f988a12e639f1eOHpwB2N8i9ta8xko7k29ejymFwfBHRJxh+pWaTu1FkXQvn85QSXeuxBX0JI/nDe1DCUBUTnLl2Lah6/ggNGXoFlA205HsvxNzx0X69Dkjf50N1DlSdrruzjxIT/8BzYe&reflink=article_email_share
- NYC’s Deaths Mirror Patterns Elsewhere: https://www.wsj.com/articles/new-york-citys-coronavirus-deaths-match-demographics-in-other-hot-spots-11587214800?emailToken=431a69ce96c62d7963b263078ba230737i1M5JE3rrp14zRmD2CWM3JsOVaJyWmS5o0CyI69f0ZsVl8dolZ1yLPaoZtPrXy1OTsEJJYygy4rPaqPY3w2uK45f2aAFtPA007//tC6A3ZlzBkmToXjwuLlllX2LEAjTb2Pf9cH54YlNUPzTwSG2Q%3D%3D&reflink=article_email_share
- New COVID-19 Community Vulnerability Map Uses Social Determinants of Health to Identify Populations At Greater Risk: https://hitconsultant.net/2020/03/24/covid-19-community-vulnerability-map/#.XqS3EzNKhPY

## Notes

### Competing Interest Statement

The authors have declared no competing interest.

